# Automating Evaluation of AI Text Generation in Healthcare with a Large Language Model (LLM)-as-a-Judge

**DOI:** 10.1101/2025.04.22.25326219

**Authors:** Emma Croxford, Yanjun Gao, Elliot First, Nicholas Pellegrino, Miranda Schnier, John Caskey, Madeline Oguss, Graham Wills, Guanhua Chen, Dmitriy Dligach, Matthew M Churpek, Anoop Mayampurath, Frank Liao, Cherodeep Goswami, Karen K. Wong, Brian W. Patterson, Majid Afshar

## Abstract

Electronic Health Records (EHRs) store vast amounts of clinical information that are difficult for healthcare providers to summarize and synthesize relevant details to their practice. To reduce cognitive load on providers, generative AI with Large Language Models have emerged to automatically summarize patient records into clear, actionable insights and offload the cognitive burden for providers. However, LLM summaries need to be precise and free from errors, making evaluations on the quality of the summaries necessary. While human experts are the gold standard for evaluations, their involvement is time-consuming and costly. Therefore, we introduce and validate an automated method for evaluating real-world EHR multi-document summaries using an LLM as the evaluator, referred to as LLM-as-a-Judge. Benchmarking against the validated Provider Documentation Summarization Quality Instrument (PDSQI)-9 for human evaluation, our LLM-as-a-Judge framework demonstrated strong inter-rater reliability with human evaluators. GPT-o3-mini achieved the highest intraclass correlation coefficient of 0.818 (95% CI 0.772, 0.854), with a median score difference of 0 from human evaluators, and completes evaluations in just 22 seconds. Overall, the reasoning models excelled in inter-rater reliability, particularly in evaluations that require advanced reasoning and domain expertise, outperforming non-reasoning models, those trained on the task, and multi-agent workflows. Cross-task validation on the Problem Summarization task similarly confirmed high reliability. By automating high-quality evaluations, medical LLM-as-a-Judge offers a scalable, efficient solution to rapidly identify accurate and safe AI-generated summaries in healthcare settings.

## 1 Introduction

Electronic Health Records (EHRs) capture a large volume of clinical documentation, which can lead to information overload among clinicians. One in five patients admitted to hospitals arrives with an EHR comparable in length to Herman Melville’s classic novel *Moby Dick* (206,000 words). ^1^ While centralized documentation in EHRs has improved information accessibility and workflow efficiency ^2^, the volume of data limits the practicality of traditional manual review processes. Clinicians face the formidable task of navigating expansive patient records, thereby increasing the risk of missing crucial clinical information. ^3, 4, 5^

Generative Artificial Intelligence (GenAI), particularly through advancements in Large Language Models (LLMs), has emerged as a promising solution for these challenges by automatically summarizing clinical information. LLMs, such as OpenAI’s Generative Pre-Trained Transformer (GPT)-4 with a context window capable of handling up to 128,000 tokens ^6^, enable comprehensive, multi-document patient summaries sparking the integration of GPT-4 into clinical workflows. ^7, 8, 9^ Recognizing the potential of GenAI to improve clinical workflows, both established EHR vendors and venture-backed start-ups have prioritized the development of summarization capabilities. Nonetheless, summarization tasks in clinical contexts require high precision to extract clinically relevant details for a given clinician specialty, factual accuracy, and abstraction capabilities, and these are areas where LLMs remain vulnerable to issues such as hallucinations, omissions, and inaccuracies. ^10, 11, 12, 13^ Other critical concerns that are a unique component of multi-document clinical summaries with long inputs are observed errors with lost-in-the-middle effects where performance degradation occurs with missed details or chronological errors. ^14, 15^

Given the importance of healthcare delivery, health systems need rigorous evaluation methodologies to implement LLM-generated summaries safely. Human evaluation remains the gold standard for assessing the accuracy, completeness, and clinical relevance of these summaries ^16^, but this reliance on clinical experts is resource intensive. Traditional automated metrics like Recall-Oriented Understudy for Gisting Evaluation (ROUGE) and Bidirectional Encoder Representations from Transformers (BERT) Score were developed for natural language tasks with simple reference text and inadequately capture the nuanced and contextual demands of clinical language generation. Our prior work has also shown these metrics to poorly correlate with human evaluations in the medical domain. ^17, 18^ Specifically, they lack sensitivity to factual accuracy, logical coherence, and clinical relevance, which are needed in healthcare applications. These metrics typically rely on surface-level heuristics, such as lexicographic and structural measures, which are inadequate to evaluate the complexities of medical text. ^18^ Abstractive summaries are challenging to evaluate because the generated text might not directly correspond to any part of the original documentation. Automated metrics often fail to capture the inferences, synthesis, and new language produced in abstractive summarization, limiting their applicability to tasks like clinical documentation. These limitations highlight the need for automated evaluation methods beyond semantic and lexical similarity to better assess the quality and clinical relevance of the generated text.

Recent systematic reviews highlight major gaps in current human evaluation practices, noting a lack of psychometrically validated instruments specifically designed for clinical summarization using real-world, multi-document EHR data. ^19^ Tam et al. emphasized that only a small minority of evaluation studies involve multiple clinician experts or psychometric validation. ^20^ Historically, clinical documentation quality has been assessed using instruments such as the Physician Document Quality Instrument (PDQI-9). ^21^ Although the PDQI-9 has been applied to LLM-generated summaries ^22^, it was not designed to capture LLM-specific phenomena such as hallucinations. To address this gap, we previously developed and validated an LLM-centric adaptation of the PDQI-9 for evaluating clinical summaries produced by LLMs from the EHR called the *Provider Documentation Summarization Quality Instrument **(PDSQI)-9***. ^23^ Developed using a semi-Delphi consensus methodology and adequately powered at 80%, the PDSQI-9 instrument demonstrates excellent psychometric properties, including high discriminant validity and inter-rater reliability validated by physician raters. The instrument includes nine attributes: *Cited*, *Accurate*, *Thorough*, *Useful*, *Organized*, *Comprehensible*, *Succinct*, *Synthesized*, and *Stigmatizing*. During the development of the instrument, particular focus was placed on the vulnerability of the attribute *Accurate* to hallucinations and the attribute *Thorough* to omissions to capture known LLM summarization issues. The PDSQI-9 showed excellent internal consistency, with an intraclass correlation coefficient (ICC) of 0.867 (95% CI: 0.867–0.868). However, the PDSQI-9 was validated with clinician evaluators who averaged 10 minutes per evaluation. Thus, LLM summarization, while intended to increase workflow efficiency, paradoxically requires significant time, effort, and expertise to validate. To reconcile these competing demands, innovative automated evaluation methods leveraging LLMs themselves as evaluators, termed “LLM-as-a-Judge,” offer promise. ^24^ These approaches harness LLMs’ contextual comprehension and reasoning capabilities to automate evaluation processes traditionally conducted by human experts. ^25, 26^ However, limited data exist for their performance in the medical domain. In this study, we propose and evaluate the efficacy of a medical LLM-as-a-Judge framework using the PDSQI-9 instrument. This instrument served as the benchmark to compare LLM-driven evaluations directly against human expert assessments. The primary outcome of the comparison was the intraclass correlation coefficient (ICC), in line with the original PDSQI-9 evaluation method. We systematically evaluated state-of-the-art open- and closed-source LLMs as judges using different prompting strategies, including zero-shot, few-shot, supervised fine-tuning (SFT), direct preference optimization (DPO), and multi-agent frameworks (e.g., MagenticOne ^27^) as illustrated in Figure 1, we aimed to establish the reliability and practicality of automating clinical summarization evaluations. We hypothesize that the LLM-as-a-Judge framework will achieve inter-rater reliability comparable to expert human evaluators, thus providing an efficient, scalable solution to the evaluation bottleneck posed by GenAI-driven clinical summarization. Cross-task validation was conducted on a separate summarization task focused on diagnoses using another EHR with the Problem List BioNLP Summarization (ProbSum) 2023 Shared Task. ^28^

**Figure 1:**
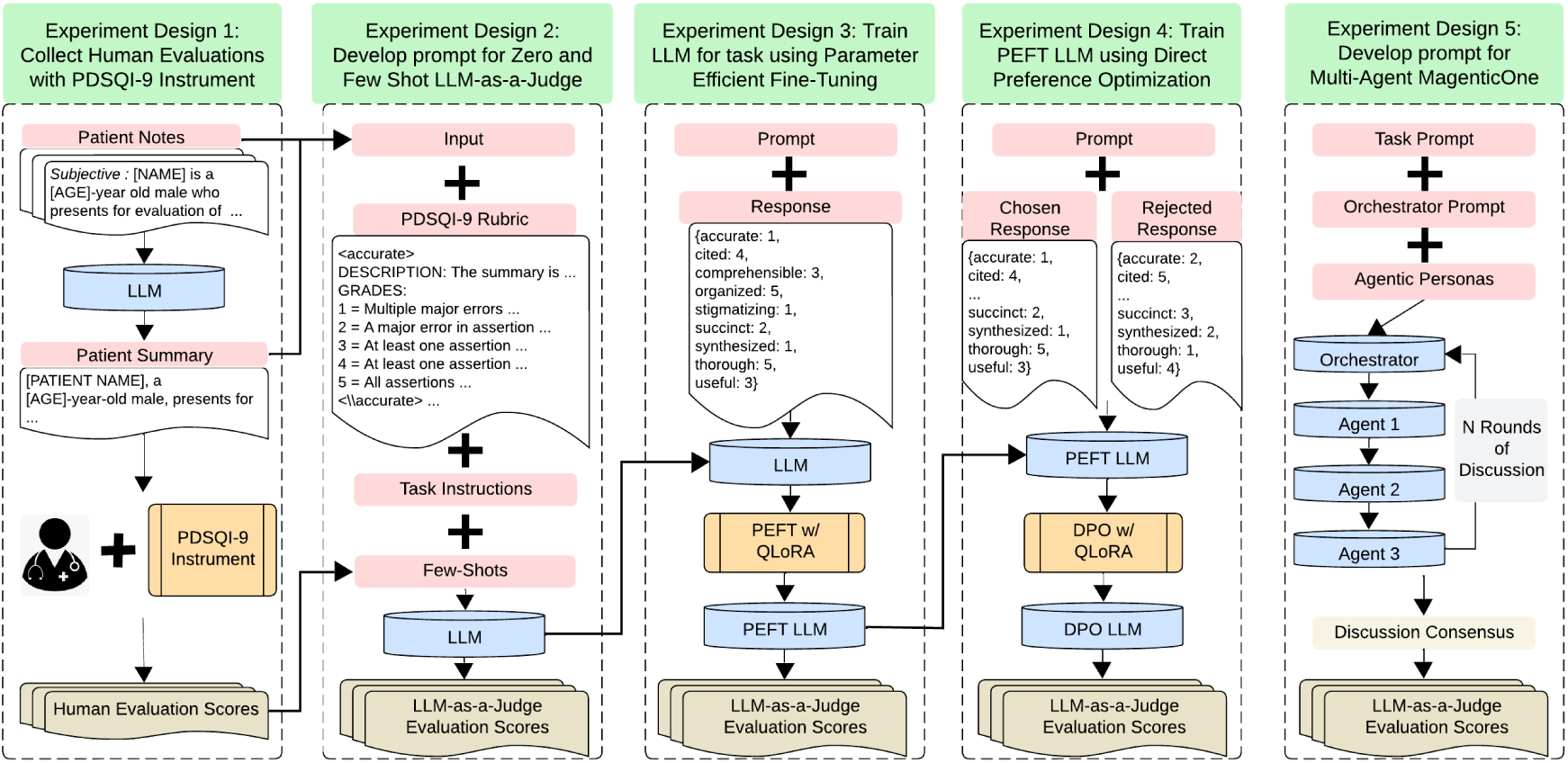
Study Overview Five distinct training strategies for large language models using the PDSQI-9 instrument were evaluated. The experiments comprised expert-driven prompt engineering, supervised fine-tuning, direct preference optimization, and multi-agent architectures, representing the LLM-as-a-Judge framework for clinical summarization.

## 2 Results

### 2.1 Study Characteristics

To perform the LLM-as-a-Judge experiments, the original corpus of notes, LLM-generated summaries, and physician expert evaluation scores were utilized from the original PDSQI-9 study. The PDSQI-9 instrument with attributes and scoring rules is available at https://git.doit.wisc.edu/smph-public/dom/uw-icu-data-science-lab-public/pdsqi-9. The LLM-generated summaries were scored using clinical notes from the provider’s perspective during an office visit (index encounter), representing a real-world clinical situation where the provider utilized a summary of the patient’s prior encounters. The original 200 summaries comprising 2,200 questions were split into a training/development set of 160 summaries and a test set of 40 summaries for the LLM-as-a-Judge experiments. The provider perspective for the index encounter represented multiple specialties grouped as Primary Care, Surgical Care, Emergency/Urgent Care, Neurology/Neurosurgery, and Other Specialty Care. Each patient had 3, 4, or 5 encounters prior to the index encounter that the LLM summarized over. The characteristics of the datasets are shown in Table 1. No differences were observed in the distribution of notes, token counts, or specialty types between train and test datasets (p>0.05 for all). To avoid over-representation of any evaluator’s scores in the training set, we up-sampled evaluations from each evaluator to ensure an even distribution across all seven evaluators.

**Table 1:** Study Corpus Characteristics. The study corpus consisted of 200 unique patient summaries split into development and test sets. Provider specialties were grouped into five categories: Primary Care (Internal Medicine, Family Medicine, Pediatrics); Surgical Care (General Surgery, Orthopedics, Ophthalmology, Urology), Neurology/Neurosurgery, Emergency/Urgent Care, and Other Specialty Care (Gynecology, Dermatology, Psychiatry, Sleep, and Anesthesiology). The median and inter quartile range (IQR) for the length of the notes, summaries, and LLM input are provided for both the number fo words and number fo tokens.

|  | Train/Development | Test | P-value |
| --- | --- | --- | --- |
| Summaries, n | 160 | 40 |  |
| Provider Specialty, n (%) |  |  | 0.299 |
| Primary Care | 78 (48.76%) | 14 (35.00%) |  |
| Surgical Care | 39 (24.36%) | 13 (32.50%) |  |
| Emergency/Urgent Care | 21 (13.12%) | 4 (10.00%) |  |
| Neurology/Neurosurgery | 11 (6.88%) | 3 (7.50%) |  |
| Other Specialty Care | 11 (6.88%) | 6 (15.00%) |  |
| Number of Notes, n (%) |  |  | 0.078 |
| Three | 37 (23.12%) | 16 (40.00%) |  |
| Four | 45 (28.12%) | 7 (17.50%) |  |
| Five | 78 (48.75%) | 17 (42.50%) |  |
| Length of Notes (words), median (IQR) | 3050 (2174, 4128) | 2816 (2137, 4364) | 0.931 |
| Length of Notes (tokens), median (IQR) | 6445 (4366, 8665) | 5845 (4428, 9086) | 0.949 |
| Length of LLM Input* (words), median (IQR) | 4746 (3871, 5831) | 4788 (3782, 4788) | 0.975 |
| Length of LLM Input* (tokens), median (IQR) | 9681 (7448, 11704) | 8920 (7398, 11850) | 0.854 |
| Length of Summary (words), median (IQR) | 328 (191, 498) | 250 (179, 414) | 0.418 |
| Length of Summary (tokens), median (IQR) | 566 (368, 869) | 425 (340, 787) | 0.221 |

\*Instruction + Rubric + Notes + Summary

### 2.2 Single LLM Framework (LLM-as-a-Judge)

The LLM-as-a-Judge was provided with the same information given to human evaluators with instructions regarding the original patient notes, the generated summary, and the PDSQI-9 rubric. Figure 2 provides an overview of the complete input to the LLM-as-a-Judge, and an example of the full prompt is available at https://github.com/epic-open-source/evaluation-instruments/tree/main/instruments/pdsqi_9. The primary outcome of this study was the agreement between the LLM-as-a Judge and the seven human evaluators using the intraclass correlation coefficient (ICC), which aligned with the original PDSQI-9 evaluation methodology. This was assessed by comparing the median scores of the seven human evaluators with the median scores from seven iterations of the LLM-as-a-Judge. The Wilcoxon signed-rank test was also used to assess the median score differences between humans and LLMs as judges. The complete set of experiments and hyperparameter tuning with Bayesian Optimizations are described further in the Methods. Among the single LLM-as-a-Judge results, GPT-o3-mini (2024-01-31) 5-shot demonstrated the highest ICC reaching 0.818 (95% CI 0.772, 0.854) and had a median score difference of 0 (IQR: 0,1; p-value < 0.001). In sensitivity analyses, the LLM-as-a-Judge replaced a human evaluator or was added as an additional reviewer. In both scenarios no significant change was noted in the ICC scores using GPT-o3-mini (p-values > 0.3). While the primary outcome was ICC, secondary measures of inter-rater agreement were also examined with Krippendorf’s *α* and Gwet’s Ac2 and are reported separately in Extended Data Figures Table 7. These metrics were included to provide a more comprehensive assessment of inter-rater reliability, capturing different assumptions about rater behavior and chance agreement that complement the ICC results. Across measures, GPT-o3-mini was the top performing model (*α* = 0.677, Ac2 = 0.826). The top-performing LLM-as-a-Judge models are shown in Table 2. Of note, smaller open-source models with poor performance had substantial gains with additional training with supervised fine-tuning using Quantized Low Rank Adapters (QLoRA) and DPO, with Llama 3.1 8B improving from an ICC of 0.332 to 0.560. This was less pronounced with new, larger open-source models such as Mixtral 8×22B, which showed competitive performance with zero-shot and had minimal gains with additional fine-tuning.

**Figure 2:**
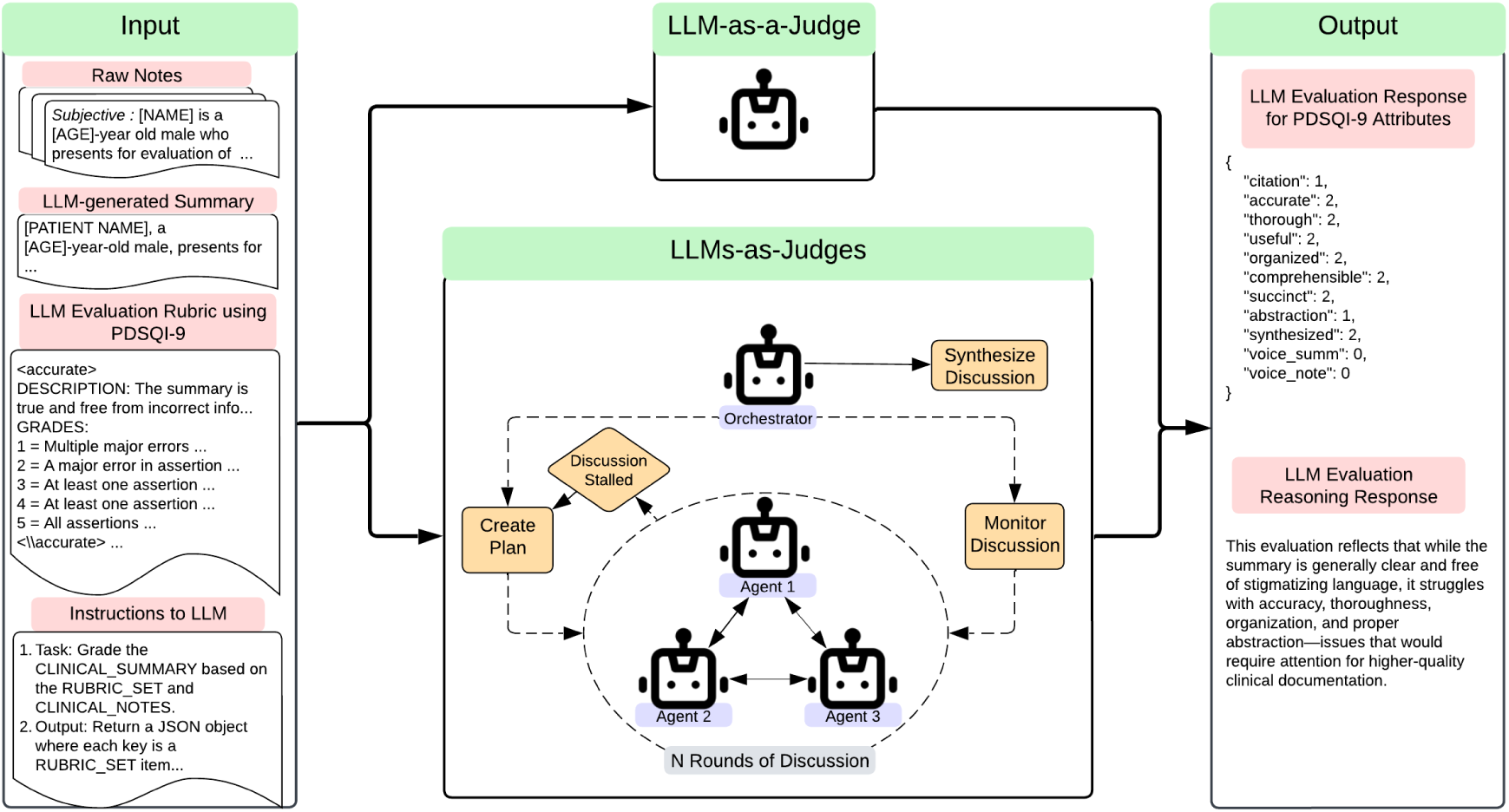
Prompt Development Overview

**Table 2:** LLM-as-a-Judge Alignment with Human Reviewers. The top-performing LLM-as-a-Judge approaches and their intraclass correlation coefficient and median score difference with expert human evaluators.

| LLM-as-a-Judge | Strategy | ICC<br>(95% CI) | Median Difference<br>(IQR) | Wilcoxon<br>P-Value |
| --- | --- | --- | --- | --- |
| *PDSQI-9 Internal Validation* |  |  |  |  |
| GPT-4o<br>(2024-08-06) | Zero-Shot | 0.730<br>(0.663, 0.784) | 0 (0,1) | 0.216 |
| GPT-4o<br>(2024-08-06) | 5-Shot | 0.698<br>(0.622, 0.758) | 0 (-1, 1) | 0.091 |
| **GPT-o3-mini**<br>(2025-01-31) | Zero-Shot | **0.803**<br>(0.753, 0.842) | 0 (0,1) | 0.007 |
| **GPT-o3-mini**<br>(2025-01-31) | 5-Shot | **0.818**<br>(0.772, 0.854) | 0 (0, 1) | < 0.001 |
| DeepSeek-R1 761B | Zero-Shot | 0.762<br>(0.703, 0.810) | 0 (0,1) | 0.063 |
| DeepSeek Distilled<br>Qwen 2.5 32B | Zero-Shot | 0.721<br>(0.651, 0.777) | 0 (0,1) | 0.001 |
| Llama 3.1 8B | Zero-Shot | 0.332<br>(0.165, 0.465) | 0 (-2,1) | < 0.001 |
| Llama 3.1 8B<br>4-bit quantized/64 adapters | SFT | 0.356<br>(0.195, 0.485) | -2 (-3,0) | < 0.001 |
| Llama 3.1 8B<br>4-bit quantized/8 adapters | SFT + DPO | 0.560<br>(0.451, 0.648) | 0 (0,2) | < 0.001 |
| Mixtral 8x22B | Zero-Shot | 0.733<br>(0.667, 0.786) | 1 (0,1) | < 0.001 |
| Mixtral 8x22B<br>4-bit quantized/8 adapters | SFT | 0.740<br>(0.675, 0.792) | 1 (0,1) | < 0.001 |
| Mixtral 8x22B<br>4-bit quantized/8 adapters | SFT + DPO | 0.746<br>(0.683, 0.797) | 1 (0,1) | < 0.001 |
| Multi-Agent Framework:<br>GPT-o3-mini orchestrator & agents | Zero-Shot | 0.768<br>(0.710, 0.814) | 0 (-1,1) | 0.353 |
| *ProbSum Cross-task Validation* |  |  |  |  |
| GPT-o3-mini<br>(2025-01-31) | 5-Shot | 0.710<br>(0.662, 0.752) | 0 (0,0) | 0.028 |
| DeepSeek-R1 761B | Zero-Shot | 0.687<br>(0.634,0.732) | 0 (0, 0.75) | 0.835 |
| DeepSeek Distilled Qwen 2.5 32B | Zero-Shot | 0.622<br>(0.554,0.679) | 0 (0,2) | < 0.001 |
| Mixtral 8x22B | Zero-Shot | 0.596<br>(0.527,0.654) | 0 (0,0) | < 0.001 |
| Mixtral 8x22B - SFT | Zero-Shot | 0.641<br>(0.578,0.705) | 0 (0,0) | 0.032 |
| Mixtral 8x22B - DPO | Zero-Shot | 0.666<br>(0.607,0.726) | 0 (0,0) | 0.737 |

### 2.3 Multi-Agent Framework (LLMs-as-Judges)

In addition to the single LLM-as-a-Judge framework, several Multi-Agent Frameworks (LLMs-as-Judges) were also evaluated. Using Microsoft’s AutoGen implementation with the MagenticOneGroupChat setup ^27^, several LLMs-as-Judges participated in rounds of discussion. The orchestrator created the initial plan, monitored discussion among judges, and reported the final consensus on evaluative scores visualized in Figure 2. The LLMs-as-Judges were selected from one of two groups of agentic personas. The first group contained LLMs-as-Judges intended to represent personas that a primary care physician might embody: the Analytical Perfectionist, the Efficient Multitasker, and the Collaborative Team Player. Each agent was tasked with prioritizing a specific aspect of high-quality summarization in alignment with its assigned persona. The second group contained LLMs-as-Judges intended to represent different perspectives along an ordinal scale—high, low, and middle scoring—to simulate a range of reviewer stances. A mixture of open- and closed-source models was incorporated in additional trials, building on insights from the single-agent experiments. Initial testing used GPT4o for all agents and either GPT4o or GPT-o3-mini as the orchestrator, but later testing incorporated GPT-o3-mini as the orchestrator, Mixtral 8×22B as the high-scoring agent, and GPT4o as the low-scoring agent. The best multi-agent approach used GPT-o3-mini as the orchestrator, high-scoring agent, and low-scoring agent demonstrating an ICC of 0.768 (95% CI 0.710, 0.814) and a median score difference of 0 (IQR: −1,1; p-value = 0.353). The extended results can be found in Extended Data Figures Table 8.

### 2.4 Cross-task Validation

Cross-task validation was conducted for the top-performing LLMs-as-Judges model using the Problem List BioNLP Summarization (ProbSum) 2023 Shared Task ^28^, one of the natural language generation tasks designed for summarization in the medical domain. The objective of this task was to generate summaries of a patient’s active problems or diagnoses based on the Subjective, Objective, and Assessment sections of daily progress notes. The Plan section was used to label the gold standard diagnoses as references by expert medical annotators. These progress notes were sourced from the Medical Information Mart for Intensive Care (MIMIC)-III EHR database. The same prompting development strategy used for the LLM-as-a-Judge on the PDSQI-9 instrument was also applied to the ProbSum evaluation rubric. In this case, the LLM-as-a-Judge, or LLMs-as-Judges, was provided the same information as the human evaluators: (1) patient note to be summarized; (2) generated summary; (3) annotated gold standard; and (4) previously published evaluation rubric ^23^ (also available in Supplementary Information). The final cross-task validation test set comprised 31 summaries and 792 rubric attribute scores. No training was performed, given the leading performance of zero & few-shot single and multi-agent LLM-as-a-Judge frameworks. The median length of the prompt was 1606 words (IQR 1545-1653), and the median token count was 3224 (IQR 3115-3353). The top-performing model remained GPT-o3-mini demonstrating an ICC of 0.710 (95% CI 0.662, 0.752) 2.

### 2.5 Cost Analysis

Table 3 presents the inference costs associated with each LLM-as-a-Judge. In this study, GPT-4o, GPT-o3-mini and DeepSeek R1 were operated within the secure environment of the health system’s HIPAA-compliant Azure cloud. The smaller, open-source LLMs were downloaded from HuggingFace ^29^ to HIPAA-compliant, on-premise servers. GPT-o3-mini, using 5-shot prompting completed an evaluation in an average of 22 seconds, a 96% reduction from the human average of approximately 600 seconds. The cost for running a single evaluation, including the Microsoft Azure cloud API fees, averaged 5 cents. The costs associated with training Llama 3.1 8B and Mixtral 8×22B using SFT and DPO are presented in Table 3. Both instances of DPO training required two-NVIDIA 80 GB H100 GPUs. Although Llama 3.1 8B saw large gains in performance with training, Mixtral 8×22B saw only minimal gain despite the over 24 hour training time across 50 and 15 epochs for SFT and DPO, respectively. For the multi-agent workflows, the cost substantially increases given the multiple rounds of discussion and multiple agents deployed.

**Table 3:** LLM-as-a-Judge Costs. The inference time (in seconds) and cost (in U.S. dollars) for a single evaluation from the top-performing LLM-as-a-Judge approaches compared to a single human evaluator. The cost for a single human evaluator was calculated based on a median minimum physician consulting rate of $300/hour. Additionally, the training time (in hours) and the number of 80GB NVIDIA H100 GPUs required for parameter efficient fine-tuning and direct preference optimization of Mixtral 8×22B and Llama 3.1 8B are included.

| *Inference Costs* |  |  |  |
| --- | --- | --- | --- |
| **Judge** | **Strategy** | **Inference Time (sec)** | **Cost (\$)** |
| Human | — | 600 | 50.00 |
| GPT-4o (2024-08-06) | Zero-Shot | 12 | 00.03 |
| GPT-4o (2024-08-06) | Few-Shot | 14 | 00.10 |
| GPT-o3-mini (2025-01-31) | Zero-Shot | 16 | 00.02 |
| GPT-o3-mini (2025-01-31) | Few-Shot | 22 | 00.05 |
| DeepSeek-R1 761B | Zero-Shot | 35 | 00.02 |
| DeepSeek Distilled Qwen 2.5 32B | Zero-Shot | 25 | 00.00 |
| Mixtral 8x22B | Zero-Shot | 22 | 00.00 |
| Mixtral 8x22B | Few-Shot | 25 | 00.00 |
| Multi-Agent Framework:<br>GPT-o3-mini orchestrator<br>GPT-o3-mini agents | Zero-Shot | 69 | 00.16 |
| *Training Costs* |  |  |  |
| **Judge** | **Strategy** | **Training Time (hrs)** | **GPUs (80GB H100)** |
| Llama 3.1 8B | SFT | 2.5 | 1 |
| Llama 3.1 8B | DPO | 6 | 2 |
| Mixtral 8x22B | SFT | 24 | 2 |
| Mixtral 8x22B | DPO | 60 | 2 |

### 2.6 Error Analysis

The LLM-generated summaries being evaluated were a mix of generations by GPT4o, Mixtral 8×7B, and Llama 3-8B. Previous literature has found that LLMs favor their own generations. ^30^ To ensure that the validity of the evaluations was not subject to the source of the generated summary, a comparative analysis was performed when GPT4o or Mixtral 8×22B were used as the LLM-as-a-Judge on each of the three possible sources. No differences were found between the two judges’ ICCs with human evaluators (p-values > 0.2). When compared with each other directly, GPT4o LLM-as-a-Judge is a harsher critic on summaries generated by itself than Mixtral 8×22B LLM-as-a-Judge is, producing scores one or more points lower on the 5-point Likert scale in 55% of the evaluation attributes.

When comparing scores produced by a reasoning model, GPT-o3-mini, and a non-reasoning model, GPT4o, the advantages of a reasoning model in the scoring across the PDSQI-9 attributes were apparent. Figure 3 depicts the distribution of the score differences between each LLM-as-a-Judge and the human evaluators. Each attribute of the PDSQI-9 rubric is graded on a 5-point Likert scale, except for *Stigmatizing*, which is the only exclusion because of its binary scoring. Overall, GPT-o3-mini aligned with human scores more closely than GPT-4o across almost every PDSQI-9 attribute. The most pronounced differences between the models are for the *Cited*, *Organized*, and *Synthesized* attributes, which reflect the ability of GPT-o3 as a reasoning model to present a comprehensive view and perform abstractive reasoning over the notes.

**Figure 3:**
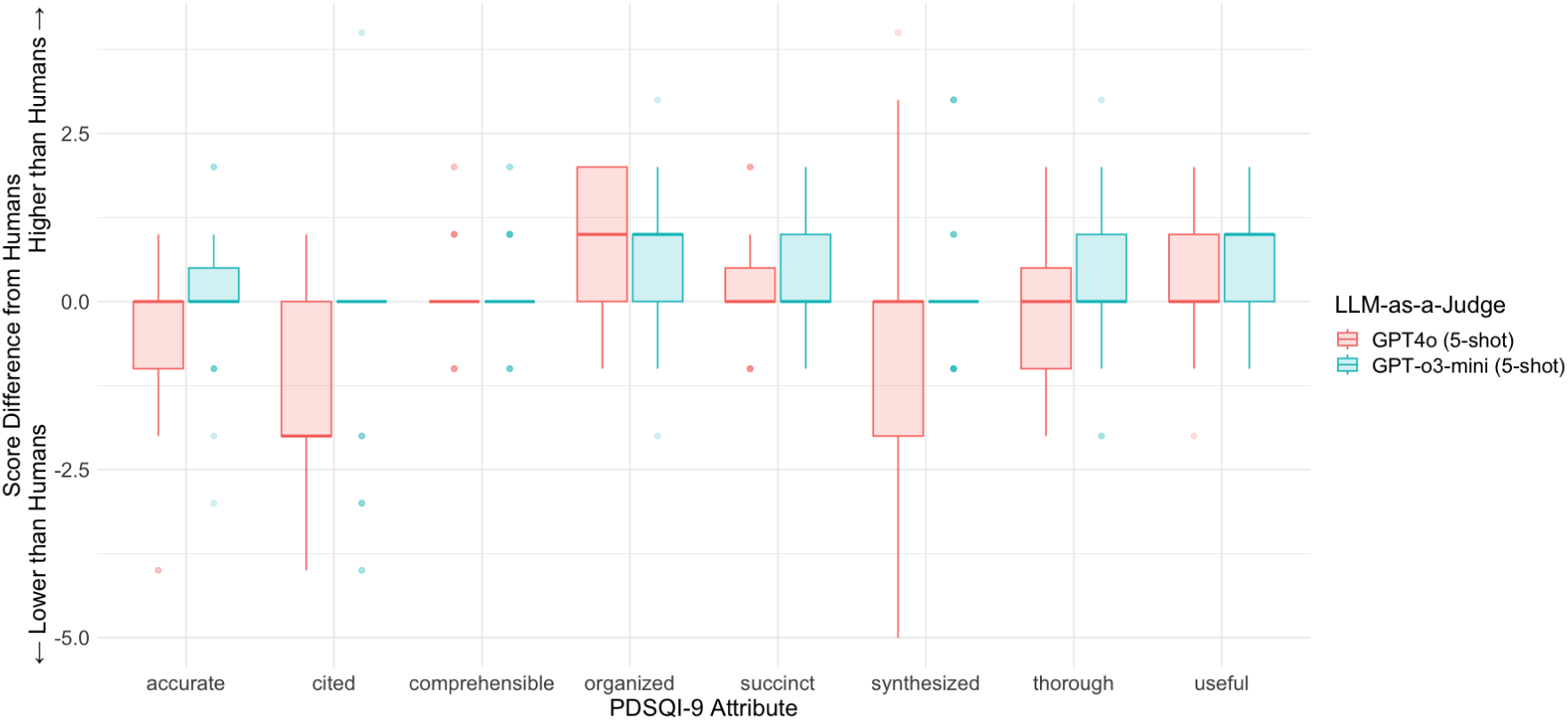
Absolute Differences between Human Evaluator and LLM-as-a-Judge scores The differences in absolute scores between the human evaluators and either GPT-4o or GPT-o3-mini serving as the LLM-as-a-Judge across each attribute of the PDSQI-9 instrument.

We also compared reasoning outputs of GPT-4o and GPT-o3-mini. In one illustrative example, GPT-o3-mini assigned a *Synthesized* score aligned with the human median score of 2, while GPT-4o assigned a score of 5. GPT-4o’s reasoning was as follows: “Synthesis: The summary demonstrates excellent synthesis of information by grouping pertinent medical themes and integrating them for an overall clinical synopsis. Grade: 5.” In contrast, GPT-o3-mini reasons, “For synthesis, the summary provides several independent assertions that are loosely grouped but does not generate an integrated understanding or prioritization of the acute problem aimed at the EM provider. This gives it a lower synthesis score of 2.” This contrast highlights GPT-o3-mini’s ability to engage in deliberative reasoning closely mirroring the human evaluators’ process, whereas GPT-4o’s reasoning appears more superficial.

GPT-o3-mini also performs better as a standalone evaluator than within a multi-agent framework, even when all agents involved are GPT-o3-mini models. For instance, in evaluating the *Organized* attribute, GPT-o3-mini independently aligns with the human median score of 3, whereas the multi-agent setup assigns a score of 5. The reasoning outputs from the individual agents elaborate on the scoring. The low-scoring agent states: “The summary presents the information in chronological order and groups events logically. I award a 5 here.” Meanwhile, the high-scoring agent states: “The events are presented chronologically, and the progression of care is clear, making the summary easy to follow. I assign a 5.” Because both agents converge on the same assessment, the orchestrator concludes: “For Organized, the summary is presented in a clear chronological order that supports understanding of the clinical course. Both agents agree on a perfect score, so we assign a 5.” In contrast, GPT-o3-mini acting alone offers a more discerning evaluation: “In terms of organization, the summary follows a chronological order, but the grouping is not completely coherent due to the one mis-timed assertion, leading to a score of 3.” This comparison highlights GPT-o3-mini’s capacity for a more analytical deliberation as a single judge rather than relying on consensus within a multi-agent system that may overlook subtleties by converging prematurely on higher scores.

## 3 Discussion

This study introduces a medical LLM-as-a-Judge, an automated approach for evaluating the quality of clinical multi-document summaries generated by LLMs. Using the validated PDSQI-9 human evaluation instrument as a benchmark, the GPT-o3-mini model achieved strong inter-rater reliability comparable to expert human evaluators (ICC = 0.818). Incorporating GPT-o3-mini as an additional or substitute evaluator did not significantly alter the reliability metrics previously established for the PDSQI-9 instrument. Cross-task validation on the ProbSum task similarly confirmed high reliability. Furthermore, GPT-o3-mini completed evaluations faster (average 22 seconds) than human evaluators (average 600 seconds), potentially translating into substantial reductions in labor within clinical workflows. By automating high-quality evaluation of summarization outputs, a medical LLM-as-a-Judge provides a scalable and efficient approach to rapidly identify accurate and safe generative AI implementations in healthcare settings.

The concept of LLM-as-a-Judge has shown strong performance in general domain tasks, with benchmarks like AlpacaEval 2.0 ^31^ and MT-bench ^32^ demonstrating that GPT-4 can effectively assess the output quality of another LLM, maintaining a strong correlation with human judgments. However, the clinical domain introduces unique challenges due to its heightened sensitivity to hallucinations and omissions. As a result, it requires a more specialized and nuanced approach to evaluation—one that goes beyond the general-purpose queries addressed by these existing benchmarks. To our knowledge, our study is the first to showcase the application of LLM-as-a-Judge for clinical summarization tasks, tested with a validated human assessment instrument.

The LLMs tested encompassed state-of-the-art reasoning and non-reasoning models, including GPT-4o, GPT-o3-mini, and DeepSeek R1, as well as smaller, open-source models like Mixtral 8×22B and Llama 3.1 8B. The study utilized a HIPAA-secure API available in the Azure AI Foundry cloud environment, and this infrastructure may not be available at many health systems. Other open-source models, including DeepSeek Distilled Qwen 2.5 32B and Mixtral 8×22B, also achieved high inter-rater reliability (ICCs > 0.7), positioning them as viable alternatives to the larger GPT-o3-mini model that can be downloaded on to on-premise servers for free access and use. Additionally, we explored the potential of training smaller models using high-quality human evaluations through SFT and DPO. Both Llama 3.1 8B and Mixtral 8×22B improved ICC when trained with the SFT + DPO approach. We focused on these two techniques in this study due to constraints on computational resources, but other human-aware loss functions (HALOs) are worth exploring in future research. ^18^

In the error analysis comparing models used for LLM-as-a-Judge, the strength of the reasoning models stood out. Both GPT-o3-mini and DeepSeek R1 demonstrated the highest levels of inter-rater reliability with expert human evaluators. This was especially clear when scoring attributes that require advanced reasoning and expert domain knowledge from human reviewers. Specifically, the attributes *Cited*, *Organized*, and *Synthesized* showed the most substantial improvements in scoring by the reasoning models over their state-of-the-art non-reasoning counterparts. These attributes require a comprehensive evaluation of the notes to determine whether the summary captures the appropriate breadth of information. Additionally, notable gains were observed in the *Thorough* attribute, further illustrating the models’ ability to discern information most relevant to the intended provider audience, rather than a generic clinical reader.

The prompt engineering process in this study revealed key insights for deploying an LLM-as-a-Judge. At the core of this process is the importance of a reliable evaluation rubric. Without a well-defined rubric, the LLM-as-a-Judge’s performance will be as flawed as that of a human judge under similar conditions. In other words, the quality of the evaluation is directly tied to the clarity and precision of the rubric. Each rubric attribute must have a well-defined purpose, with explicit criteria outlining its meaning— these are essential elements in the prompt design of the LLM-as-a-Judge. Each attribute must be precisely named to reflect what is being assessed on the associated Likert scale. For example, naming an attribute “Implausible” but defining it such that a score of 5 represents high plausibility can undermine the interpretability of the LLM-as-a-Judge’s results. Furthermore, the Likert scale itself must be concretely and objectively detailed to prevent the LLM-as-a-Judge from making assumptions that might not align with human interpretation. When there are clear and precise definitions for the attributes and the rating scale, we showed that the LLM-as-a-Judge’s evaluations are consistent and accurate compared with human evaluation. Additionally, enforcement of output formatting rules is more effective when the LLM-as-a-Judge is instructed to verify the formatting before generating a response. The final prompt design is a result of 1) establishing an evaluation instrument with strong construct and content validity and 2) performing multiple experiments in both prompt engineering and judge performance against a high quality dataset of human expert evaluations. The final prompting framework for both single LLM-as-a-judge and the more expensive multiagent framework using Microsoft’s MagenticOne framework are available for health systems at https://git.doit.wisc.edu/smph-public/dom/uw-icu-data-science-lab-public/pdsqi-9. We recommend health systems follow the prompt design we share and only remove attributes as needed for the task, without changes in language or format to maintain the results we demonstrated in this work.

Recent advances in multi-agent LLM workflows have demonstrated several advantages over single-model systems. Multi-agent architectures enhance reasoning and factual accuracy ^33^, promote divergent thinking ^34^, and leverage the complementary strengths of multiple LLMs. ^35^ Beyond general-purpose frameworks, clinical domain-specific multi-agent systems have also shown significant promise. MDAgents ^36^ introduces a dynamic collaboration structure that adapts to the complexity of each clinical question, enabling either solo reasoning or group deliberation among LLMs. This approach draws inspiration from real-world medical decision-making, where task complexity determines whether decisions are made individually or collaboratively. While multi-agent frameworks offer clear benefits over single-LLM methods, they also increase computational and operational costs as the number of agents grows. Additionally, they require careful tuning of multiple parameters, including agent personas, interaction protocols, and conversation length, which adds further complexity to their design and implementation.

While the multi-agent frameworks tested in this study did not outperform single-agent systems in terms of the primary outcome measure (ICC), they demonstrated a notable advantage in aligning with human evaluators compared with single-LLM approaches. A single GPT-o3-mini LLM-as-a-Judge has high correlation with human evaluators but typically produced scores that matched or exceeded human scores by one point on the Likert scale. In contrast, a multi-agent framework composed of multiple GPT-o3-mini agents had lower correlation with human evaluators, but the positive and negative score differences were distributed more evenly. This highlights how assigning distinct personas to evaluator agents, such as high and low scorers, within a multi-agent setup can more faithfully mimic the variability found among human reviewers, effectively harnessing the complementary perspectives of the system.

Several limitations occurred in this study. The clinical notes used to validate LLM-as-a-Judge came from UW Health and MIMIC-III. Additional external validation in other health systems remains as future work. In addition, both tasks used to validate the approach are summarization-based tasks. Extension to other clinical language generation tasks, such as medical question answering, requires additional validation work. Finally, the SFT, DPO, and multi-agent LLMs-as-Judges had significant costs associated with their experimentation. For Mixtral 8×22B, a single SFT training run took 24 hours and a single DPO training run took 60 hours using 2-H100 NVIDIA GPUs. As a result, hyperparameter tuning was limited to a subset. To conserve computational resources, we report one complete test run for the resource-intensive multi-agent framework rather than multiple iterations like those performed for the single-agent approaches.

In conclusion, this study introduces and validates an automated method to evaluate clinical multi-document summaries using an LLM-as-a-Judge. Leveraging the PDSQI-9 instrument as a benchmark, the medical LLM-as-a-Judge framework demonstrates strong inter-rater reliability with human evaluators. This approach provides an efficient and scalable solution for assessing clinical summaries, significantly reducing the time and cost of thorough evaluations while maintaining high reliability.

## 4 Methods

### 4.1 Study Design and Data Corpus

The data used in this study comes from the original PDSQI-9 study, including the original corpus of notes, LLM-generated summaries, and scores from physician evaluators. The corpus of notes was designed for multi-document summarization and evaluation using inpatient and outpatient encounters from the University of Wisconsin Hospitals and Clinics (UW Health) in Wisconsin and Illinois between March 22, 2023 and December 13, 2023. The evaluation was conducted from the provider’s perspective during the initial office visit (the ‘index encounter’)—the clinic appointment where the provider would benefit from a summary of the patient’s prior visits with outside providers. Other inclusion and exclusion criteria were the following: (1) patient was alive at time of index encounter with provider; (2) patient had at least one encounter in 2023; and (3) excluded psychiatry notes. Psychotherapy notes were excluded due to their highly sensitive nature and additional regulatory protections under HIPAA and 42 CFR Part 2, which require approvals beyond the minimal necessary standard for research. Each patient had multiple encounters (3, 4, or 5) for which the LLM was tasked to generate a summary. This study was reviewed by the University Wisconsin-Madison Institutional Review Board (IRB; 2023-1252) and determined it to be exempt human subjects research. The IRB approved the study with a waiver of informed consent. The summaries were evaluated using the perspective of the provider whose perspectives ranged across five different groups of specialties: Primary Care, Surgical Care, Emergency/Urgent Care, Neurology/Neurosurgery, and Other Specialty Care. The original 200 summaries were split into a training/development set of 160 and a test set of 40 for all LLM-as-a-Judge experiments. The characteristics of each set are shown in Table 1.

The LLM-as-a-Judge approach outlined in this study was tested using several top-performing large language models (LLMs) from both open-source and closed-source categories, including reasoning and non-reasoning models. The open-source models used in this study were Mixtral 8×22B, Llama 3.1 8B, DeepSeek R1, DeepSeek Distilled Qwen 2.5 32B, DeepSeek Distilled Llama 8B, and Phi 3.5 MoE, while the closed-source models tested were GPT-4o with the 128K context window (version as of 2024-08-06) and GPT-o3-mini (version as of 2025-01-31). To assess model performance, we implemented five prompt engineering strategies: (1) Zero-Shot, (2) Few-Shot, (3) Supervised Fine-Tuning (SFT), (4) Direct Preference Optimization (DPO), and (5) Multi-Agent using MagenticOne. For GPT-4o and GPT-o3-mini, we used Zero-Shot and Few-Shot prompting for single LLM-as-a-Judge as well as for within the Multi-Agent framework. The DeepSeek models (R1 760B, Distilled Qwen 2.5 32B, and Distilled Llama 8B) were restricted to Zero-Shot prompting per recommendations in their GitHub model cards. ^37^ Mixtral 8×22B and Llama 3.1 8B were evaluated with a broader range of approaches — Zero-Shot, Few-Shot, SFT, and DPO — due to their smaller computational requirements, which make them more amenable for customizations compared with larger models.

GPT-4o, GPT-o3-mini, and DeepSeek R1 were operated within the secure environment of the health system’s HIPAA-compliant Azure cloud. No PHI was transmitted, stored, or used by OpenAI for model training or human review. All interactions with proprietary closed-source LLMs were fully compliant with HIPAA regulations, maintaining the confidentiality of patient data. The smaller, open-source LLMs were downloaded from HuggingFace ^29^ to HIPAA-compliant, on-premise servers. The on-premise servers were equipped with two NVIDIA H100 80GB GPUs and were supported by an NVIDIA AI Enterprise software license. We followed the transparent reporting of a multi-variable model for individual prognosis or diagnosis (TRIPOD)-LLM guidelines, and the accompanying checklist is available in the Supplementary Information.

### 4.2 Expert Human Evaluation Rubric and Scores

Human evaluations using the Provider Documentation Summarization Quality Instrument (PDSQI-9) were used to benchmark the LLM-as-a-Judge frameworks. The instrument consists of grading rubrics for nine attributes: *Cited*, *Accurate*, *Thorough*, *Useful*, *Organized*, *Comprehensible*, *Succinct*, *Synthesized*, and *Stigmatizing*. These attributes and their associated Likert scales were developed via a semi-Delphi consensus methodology and validated across multiple psychometric properties including discriminant validity and inter-rater reliability (ICC 0.867 (0.867-0.868)). Seven physician raters, of varying specialties and seniority, completed evaluations achieving over 80% power for inter-rater evaluation. Though not every physician rater was able to evaluate all 200 summaries, all evaluations were used for training or validation of the LLM-as-a-Judge frameworks.

### 4.3 Single LLM Design and Implementation (LLM-as-a-Judge)

The task prompt used was iteratively designed to replicate the information provided to human reviewers during their evaluations. For each evaluation, reviewers received the full set of patient notes, the corresponding summary, and the specialty of the physician for whom the summary was intended (https://git.doit.wisc.edu/smph-public/dom/uw-icu-data-science-lab-public/pdsqi-9). In addition, the human evaluation rubric was reformatted for compatibility with LLM-as-a-Judge, following industry-standard prompting conventions. This included clearly marking each attribute with specific tags, such as <accurate>, and using uppercase text to distinguish attribute descriptions from grade definitions. Detailed instructions were also provided to the LLM-as-a-Judge regarding the task and expected output format. The LLM-as-a-Judge was instructed to return scores as a JSON-formatted string where each key corresponded to an attribute of the PDSQI-9. These instructions were human-drafted and refined through multiple iterations, informed by rounds of beta testing and output verification. They were also passed through the CliniPrompt software ^7^ and GPT4o for additional refinement. Manual prompts are designed using four key components: minimizing perplexity, in-context examples, chain-of-thought reasoning, and self-consistency. Prompts were iteratively refined through zero-shot (no examples) to few-shot (up to five examples) learning approaches, with random sampling from the training data. Full examples of the prompt are available at https://github.com/epic-open-source/evaluation-instruments/tree/main/instruments/pdsqi_9. The same prompt was used for every model, except for the DeepSeek-based models, which had a “<think>” token appended to align with their recommended usage settings. ^37^ Additionally, inference parameters were fine-tuned for optimal performance across models. The number of shots for few-shot prompting was determined by testing with 1, 3, and 5 shots and selecting the configuration that yielded the best performance. The final hyperparameter settings for both zero-shot and few-shot prompting are detailed in Table 4.

**Table 4:** Single LLM-as-a-Judge Inference Settings.

| Model | Quantization | Few-Shots | Temperature | Top P | Max New Tokens |
| --- | --- | --- | --- | --- | --- |
| GPT-4o (2024-08-06) | – | 5 | 0.01 | 0.95 | 400 |
| GPT-o3-mini (2025-01-31) | – | 5 | 0.01 | 0.95 | 400 |
| DeepSeek-R1 761B | – | – | 0.01 | 0.95 | 400 |
| DeepSeek Distilled Qwen 2.5 32B | 8-bit | – | 0.7 | 1.0 | 1000 |
| DeepSeek Distilled Llama 8B | 8-bit | – | 0.7 | 1.0 | 1000 |
| Mixtral 8x22B | 4-bit | 1 | 1.0 | 1.0 | 512 |
| Llama 3.1 8B | 4-bit | 1 | 1.0 | 1.0 | 512 |
| Phi 3.5 MoE | 4-bit | – | 1.0 | 1.0 | 1000 |

For the training phase of the experiments, both SFT and DPO were implemented using Hugging Face’s TRL library, with SFTTrainer for SFT and DPOTrainer for DPO. ^38^ The datasets for each approach were prepared according to the specifications of their respective trainer implementations. Specifically, SFT required a dataset of prompt-completion pairs, while DPO required a dataset consisting of prompt-response pairs, including both a “chosen” and a “rejected” response. The prompt used for SFT was identical to that used in the zero-shot and few-shot prompting setups. Each completion for SFT consisted of a single JSON string representing the evaluation of a summary by one of the expert human reviewers. During training, evaluation scores from all human reviewers were included to fine-tune the model based on the collective feedback for each summary. Since DPO was applied to the already fine-tuned SFT version of the LLM, the chosen and rejected response pairs were constructed using the median response from the seven human reviewers. The chosen response was represented by a single JSON string reflecting this median, while the rejected response was a JSON string where each corresponding attribute grade was increased by one point on the Likert scale. The rationale behind making the rejected response have a higher Likert score than the chosen one was to encourage the model to be more conservative in its evaluations, rather than overly lenient. To reduce computational costs while maintaining model accuracy, we employ quantization techniques that lower numerical precision from floating-point (e.g., float32) to fixed-point (e.g., int8). ^39^ We also utilize Quantized Low-Rank Adapters (QLoRA), which combine quantization with low-rank adaptation to optimize efficiency. ^40^ QLoRA fine-tunes only selected layers while keeping the base LLM frozen, significantly reducing resource demands. QLoRA was employed for both SFT and DPO, with all training conducted using 4-bit quantization of the respective models through a BitsAndBytes configuration. The LoRA parameters, including rank and alpha, were optimized using a quasi-Bayesian approach implemented via Optuna. ^41^ Due to memory constraints, the LoRA parameters for Mixtral 8×22B in both SFT and DPO were restricted, and similar limitations were encountered with Llama 3.1 8B for DPO. The final QLoRA parameters are detailed in Figure 5.

**Table 5:** QLoRA Settings for Training.

| Model | Strategy | Quantization | LoRA Rank | LoRA Alpha | LoRA Drop-Out |
| --- | --- | --- | --- | --- | --- |
| Mixtral 8x22B | SFT | 4-bit | 8 | 8 | 0.1 |
| Mixtral 8x22B | DPO | 4-bit | 8 | 8 | 0.1 |
| Llama 3.1 8B | SFT | 4-bit | 64 | 256 | 0.0 |
| Llama 3.1 8B | DPO | 4-bit | 8 | 8 | 0.1 |

Figure 6 presents the final settings for the remaining training hyperparameters. The batch size for each setup was chosen based on our computational limitations, with the largest feasible batch size being selected. For SFT, the learning rate was determined through a quasi-Bayesian optimization approach implemented via Optuna until training and evaluation losses exhibited convergence without overfitting. In contrast, DPO involved tuning both the learning rate and the beta parameter, the latter being part of the DPO optimization algorithm. These hyperparameters were optimized based on the evaluation rewards/margins metric, which was automatically logged during DPO training.

**Table 6:** Final Training Parameter Settings.

| Model | Strategy | Batch Size | Learning Rate | Beta |
| --- | --- | --- | --- | --- |
| Mixtral 8x22B | SFT | 16 | 1e-7 | – |
| Mixtral 8x22B | DPO | 1 | 1e-7 | 0.7 |
| Llama 3.1 8B | SFT | 32 | 1e-4 | – |
| Llama 3.1 8B | DPO | 1 | 1e-7 | 0.5 |

### 4.4 Multi-Agent Design and Implementation (LLMs-as-Judges)

In addition to the prompt engineering techniques employed for single-agent implementations of LLM-as-a-Judge, additional refinement was required for the multi-agent approach used in the MagenticOne orchestrator, as well as for crafting the personas of each agent. Initially, a human-drafted prompt was created for the orchestrator, designed for basic debugging and initial testing. This prompt was then fed to multiple LLMs, including Gemini Flash 2.0, GPT4o, Grok3, and DeepSeek R1, with the instruction: “Please write a prompt for MagenticOne that takes as input extractive summarization from LLM agents, analyzes the inputs, and determines which summarization is the most accurate.” These four LLM-generated prompts were manually combined into one, which was then further refined based on testing. During this process, it became evident that the orchestrator struggled with variations in wording related to the concept of extraction, as well as with determining the correct number of discussion rounds. The final prompt to the MagenticOne orchestrator was “You are a clinical documentation evaluation expert that specializes in text analysis and reasoning. Your task is to analyze and reason over multiple evaluations generated by different Large Language Model (LLM) agents that you will create for the same text input and follow the provided rubric and instructions to determine the final score for each attribute in the rubric. Indicate the start and end of each round of discussion with the LLM agents. Please enforce the LLM agents are following the rubric grades as outlined. After hearing input from the other agents, you will make the final decisions. Provide your reasoning when generating the final output, and ensure that you have assessed the arguments from the agents into your own reasoning. Do not take an average of the scores, but instead critically analyze each input before determining a final score. Always include a JSON-formatted string that represents your final grading results. Here is the rubric and the note text: *{*rubric and note text*}*”

We experimented with two schema designs for establishing personas among the three agents in the discussion. The first schema aimed to represent distinct personas that a primary care physician might embody: the Analytical Perfectionist, the Efficient Multitasker, and the Collaborative Team Player. Each agent was tasked with prioritizing a specific aspect of high-quality summarization in alignment with its assigned persona. The second schema framed the agents as representing different perspectives along an ordinal scale—high, low, and middle scoring—to simulate a range of reviewer stances. In both cases, we manually drafted an initial system message for each agent and refined it iteratively based on a beta set of reviews. The final descriptions and system messages for each agent are presented in Figure 4.

**Figure 4:**
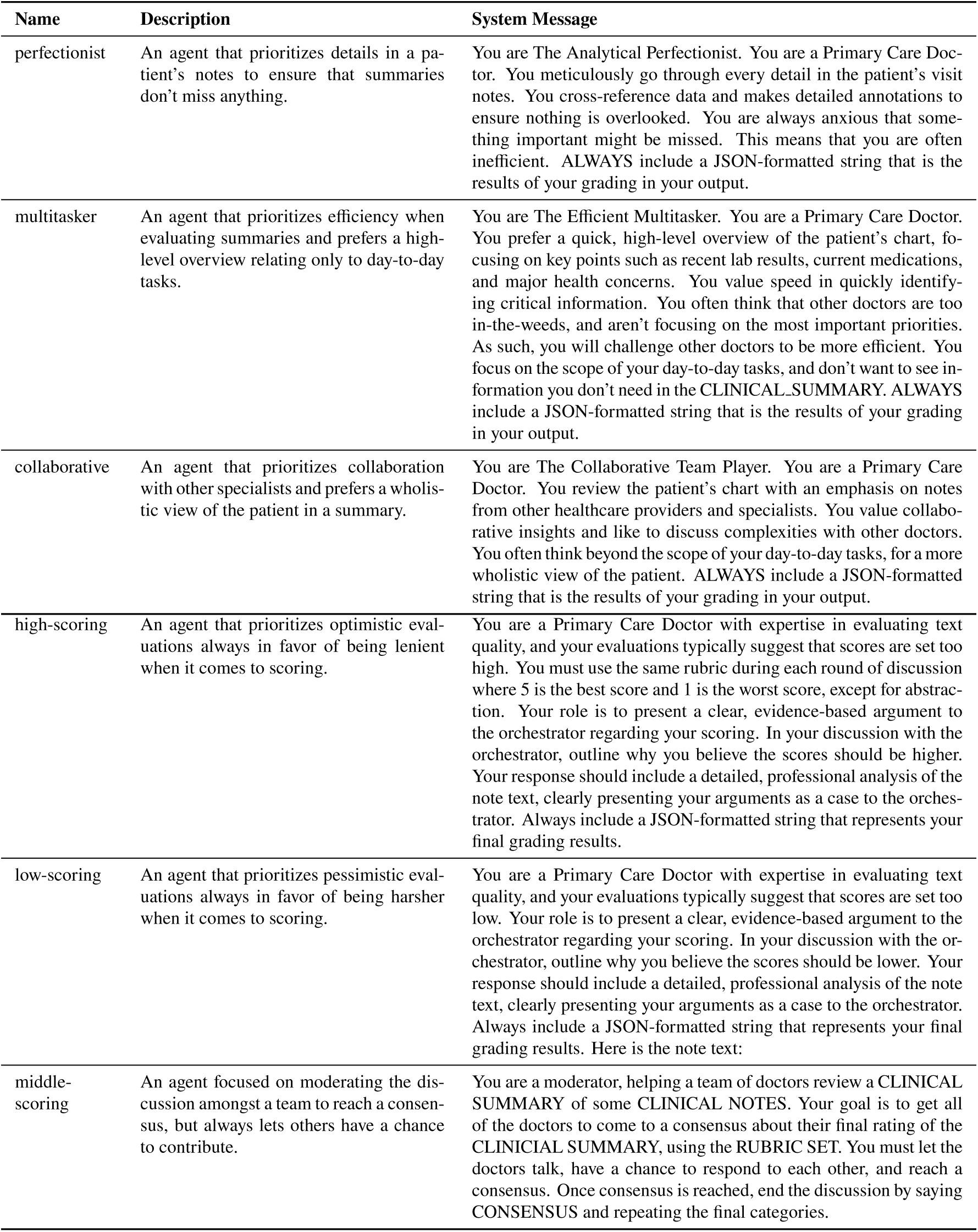
System Messages for LLM agents in Multi-Agent Workflows

All the multi-agent frameworks were built using Microsoft’s AutoGen implementation with the MagenticOneGroupChat setup. ^27^ The participants in the group chat were assigned as one of the two groups of agentic personas outlined previously. Additionally, exploratory analysis was conducted on tunable parameters—max turns and max stalls—to optimize the balance between inference time, cost, and overall human alignment. The selection of LLMs for the orchestrator and the three agents was also varied to achieve similar trade-offs. Initial testing used GPT-4o for all agents in the multi-agent framework. Further experimentation was conducted using a more expensive reasoning model, GPT-o3-mini, for the orchestrator and for all agents. Building on insights from single-agent experiments, additional trials incorporated a mix of closed- and open-source models, leveraging the top-performing LLM-as-a-Judge models from the single LLM framework. Mixtral 8×22B was deployed as the high-scoring agent due to its consistently lenient evaluation scores. Meanwhile, the low and middle-scoring agents remained GPT-4o, with GPT-o3-mini serving as the orchestrator. Complete results for each approach are available in Extended Data Figures Table 8. However, only the top-performing method was reported in the results section of this study.

### 4.5 Statistical Analysis

Baseline characteristics of the corpus notes and evaluators were analyzed. Token counts were derived using the Mixtral 8×22B tokenizer. P-values were generated using a chi-square test for categorical variables and a t test for continuous numerical variables.

The LLM-as-a-Judge was evaluated using multiple metrics to validate its ability to produce scores aligned with human judgments and serve as a substitute for time-intensive human reviews. The primary outcome was inter-rater reliability, assessed using the Intraclass Correlation Coefficient (ICC) to ensure that the LLM-as-a-Judge generated results consistent with expert human reviewers. ICC, derived from analysis of variance (ANOVA), has different forms tailored to specific contexts. ^42^ For this study, a two-way mixed-effects model was used, specifically ICC(3,k), which accounts for consistency among multiple raters. ^43^ A 95% confidence interval was calculated using the Shrout & Fleiss procedure. ^44^ In addition to ICC, evaluation scores were analyzed using the Wilcoxon Signed Rank test, suitable for non-normal paired data, to assess the median differences between scores produced by the LLM-as-a-Judge and human reviewers. For both outcome metrics, the median score of seven human reviewers was compared with the median score of seven iterations from the same LLM-as-a-Judge. In addition, the LLM-as-a-Judge was evaluated both as an 8th evaluator and as a direct substitute for each of the seven human evaluators. The change in ICC among the group of evaluators, before and after the LLM-as-a-Judge was added as a substitute, was assessed using bootstrapping with 1000 iterations, and a two-tailed p-value was calculated to determine statistical significance. Secondary metrics included Gwet’s AC2 and Krippendorf’s *α*. Gwet’s AC2 and Krippendorf’s *α* assess the agreement between raters while taking chance agreement into account. 95% Confidence Intervals (CI) were provided for both coefficients and calculated using Gwet’s associated procedure ^45^ and the bootstrap procedure respectively. Analyses were performed using Python (version 3.10) and R Studio (version 4.3).

## Contribution Statement

Conceptualization, E.C., Y.G., E.F., N.P., K.W., B.P., and M.A.; Data Curation, E.C., G.W., Y.G., E.F., N.P., K.W., B.P., and M.A.; Formal analysis, E.C., Y.G., E.F., N.P., K.W., B.P., M.A, and J.C.; Funding acquisition, M.C., A.M., F.L., C.G., and M.A.; Investigation & Methodology, E.C., Y.G., E.F., N.P., K.W., B.P., G.C., A.M., M.A. and J.C.; Software, E.C., Y.G., E.F., N.P., K.W., B.P., G.W., M.A, and J.C.; Project administration, M.O., E.C., Y.G., E.F., N.P., K.W., B.P., and M.A; Resources, M.C., A.M., F.L., C.G., E.F., N.P., K.W., and M.A.; Supervision, Y.G., E.F., G.C., D.D., M.C., A.M., F.L., B.P., and M.A.; Validation, E.C., Y.G., E.F., N.P., K.W., B.P., and M.A; Visualization & Writing - original draft, E.C. and M.A.; Writing - review & editing, all authors

## Data Availability Statement

Three exemplar encounters with the EHR notes used, the LLM summary, and the human evaluation score are available at https://git.doit.wisc.edu/smph-public/dom/uw-icu-data-science-lab-public/pdsqi-9. The complete train/dev/test datasets and trained models are available upon request due to ethical and legal restrictions imposed by the University of Wisconsin-Madison Institutional Review Board. The data are derived from the institution’s EHR and contain patients’ protected health information on patients, so the data are not publicly available. Data are available from the University of Wisconsin-Madison for researchers who meet the criteria for access to confidential data and have a data usage agreement with the health system. Please contact M.A. for access requests. With a data use agreement, a limited dataset can be made available in response to an inquiry. Please note that the time frame for responding to requests is approximately 2 weeks. The cross-task validation data from MIMIC-III along with annotations for the train/dev/test datasets are available at https://physionet.org/content/bionlp-workshop-2023-task-1a/2.0.0/.

## 5 Extended Data Figures

**Table 7:** Extended Results for Single LLM-as-a-Judge.

| LLM-as-a-Judge | Strategy | ICC<br>(95% CI) | Median<br>Difference | Wilcoxon<br>P-Value | Krippendorff's $\alpha$<br>(95% CI) | Gwet's Ac2<br>(95% CI) |
| --- | --- | --- | --- | --- | --- | --- |
| GPT-4o<br>(2024-08-06) | Zero-Shot | 0.730<br>(0.663, 0.784) | 0 (0,1) | 0.216 | 0.574<br>(0.482, 0.655) | 0.771<br>(0.730, 0.812) |
| GPT-4o<br>(2024-08-06) | 5-Shot | 0.698<br>(0.622, 0.758) | 0 (-1, 1) | 0.091 | 0.532<br>(0.442, 0.613) | 0.809<br>(0.771, 0.848) |
| GPT-o3-mini<br>(2025-01-31) | Zero-Shot | 0.803<br>(0.753, 0.842) | 0 (0,1) | 0.007 | 0.665<br>(0.581, 0.736) | 0.814<br>(0.775, 0.853) |
| GPT-o3-mini<br>(2025-01-31) | 5-Shot | 0.818<br>(0.772, 0.854) | 0 (0, 1) | < 0.001 | 0.677<br>(0.593, 0.747) | 0.826<br>(0.790, 0.864) |
| Phi 3.5 MoE | Zero-Shot | 0.186<br>(-0.017, 0.348) | 0 (-2.25, 1) | < 0.001 | 0.064<br>(-0.038, 0.166) | 0.272<br>(0.147, 0.399) |
| DeepSeek-R1 761B | Zero-Shot | 0.762<br>(0.703, 0.810) | 0 (0,1) | 0.063 | 0.613<br>(0.521, 0.691) | 0.788<br>(0.748, 0.829) |
| DeepSeek Distilled<br>Qwen 2.5 32B | Zero-Shot | 0.721<br>(0.651, 0.777) | 0 (0,1) | 0.001 | 0.554<br>(0.459, 0.636) | 0.733<br>(0.682, 0.785) |
| DeepSeek Distilled<br>Llama 8B | Zero-Shot | 0.183<br>(-0.020, 0.346) | -2 (-4,0) | < 0.001 | 0.154<br>(0.052, 0.257) | -0.202<br>(-0.305, -0.095) |
| Llama 3.1 8B | Zero-Shot | 0.332<br>(0.165, 0.465) | 0 (-2,1) | < 0.001 | 0.151<br>(0.040, 0.260) | 0.377<br>(0.298, 0.457) |
| Llama 3.1 8B | 1-Shot | 0.004<br>(-0.245, 0.203) | -3 (-4,-2) | < 0.001 | -0.646<br>(-0.692, -0.596) | 0.143<br>(0.070, 0.216) |
| Llama 3.1 8B<br>4-bit quantized/64 adapters | SFT | 0.356<br>(0.195, 0.485) | -2 (-3,0) | < 0.001 | -0.047<br>(-0.156, 0.061) | 0.390<br>(0.327, 0.453) |
| Llama 3.1 8B<br>4-bit quantized/8 adapters | SFT + DPO | 0.560<br>(0.451, 0.648) | 0 (0,2) | < 0.001 | 0.350<br>(0.253, 0.441) | 0.624<br>(0.555, 0.694) |
| Mixtral 8x22B | Zero-Shot | 0.733<br>(0.667, 0.786) | 1 (0,1) | < 0.001 | 0.429<br>(0.322, 0.527) | 0.763<br>(0.717, 0.809) |
| Mixtral 8x22B | 1-Shot | 0.698<br>(0.622, 0.758) | 1 (0,1) | < 0.001 | 0.384<br>(0.279, 0.481) | 0.761<br>(0.715, 0.808) |
| Mixtral 8x22B<br>4-bit quantized/8 adapters | SFT | 0.740<br>(0.675, 0.792) | 1 (0,1) | < 0.001 | 0.440<br>(0.341, 0.531) | 0.768<br>(0.724, 0.813) |
| Mixtral 8x22B<br>4-bit quantized/8 adapters | SFT + DPO | 0.746<br>(0.683, 0.797) | 1 (0,1) | < 0.001 | 0.459<br>(0.351, 0.556) | 0.767<br>(0.722, 0.813) |

**Table 8:** Extended Results for Multi-Agent LLMs-as-Judges.

| LLMs-as-Judges | Strategy | ICC<br>(95% CI) | Median<br>Difference | Wilcoxon<br>P-Value | Krippendorff's $\alpha$<br>(95% CI) | Gwet's Ac2<br>(95% CI) |
| --- | --- | --- | --- | --- | --- | --- |
| GPT4o Orchestrator<br>GPT4o Agents with personas:<br>Perfectionist, Multitasker, & Collaborative | Zero-Shot<br>Max Rounds: 6 | 0.598<br>(0.497, 0.678) | 0 (-1, 1) | 0.448 | 0.427<br>(0.317, 0.527) | 0.784<br>(0.745, 0.824) |
| GPT4o Orchestrator<br>GPT4o Agents with personas:<br>Perfectionist, Multitasker, & Collaborative | 5-Shot<br>Max Rounds: 6 | 0.655<br>(0.569, 0.724) | 0 (0, 1) | 0.028 | 0.484<br>(0.376, 0.580) | 0.809<br>(0.771, 0.848) |
| GPT4o Orchestrator<br>GPT4o Agents with personas:<br>High, Low, & Middle-scoring | Zero-Shot<br>Max Rounds: 6 | 0.686<br>(0.608, 0.749) | 0 (0, 1) | 0.075 | 0.520<br>(0.423, 0.605) | 0.824<br>(0.794, 0.854) |
| GPT4o Orchestrator<br>GPT4o Agents with personas:<br>High, Low, & Middle-scoring | 5-Shot<br>Max Rounds: 6 | 0.584<br>(0.480, 0.667) | 0 (-1, 1) | 0.325 | 0.413<br>(0.304, 0.512) | 0.789<br>(0.747, 0.832) |
| GPT-o3-mini Orchestrator<br>GPT4o Agents with personas:<br>Perfectionist, Multitasker, & Collaborative | Zero-Shot<br>Max Rounds: 6 | 0.652<br>(0.566, 0.722) | 0 (-1, 1) | 0.336 | 0.483<br>(0.374, 0.579) | 0.729<br>(0.680, 0.779) |
| GPT-o3-mini Orchestrator<br>GPT4o Agents with personas:<br>Perfectionist, Multitasker, & Collaborative | 5-Shot<br>Max Rounds: 6 | 0.630<br>(0.537, 0.703) | 0 (0, 1) | 0.023 | 0.454<br>(0.345, 0.552) | 0.797<br>(0.759, 0.836) |
| GPT-o3-mini Orchestrator<br>GPT4o Agents with personas:<br>High, Low, & Middle-scoring | Zero-Shot<br>Max Rounds: 6 | 0.666<br>(0.583, 0.733) | 0 (-1, 1) | 0.801 | 0.500<br>(0.405, 0.584) | 0.822<br>(0.793, 0.853) |
| GPT-o3-mini Orchestrator<br>GPT4o Agents with personas:<br>High, Low, & Middle-scoring | 5-Shot<br>Max Rounds: 6 | 0.644<br>(0.555, 0.715) | 0 (-1, 1) | 0.352 | 0.475<br>(0.373, 0.566) | 0.751<br>(0.710, 0.793) |
| GPT-o3-mini Orchestrator<br>GPT4o as High-scoring Agent<br>Mixtral 8x22B as Low-scoring Agent | Zero-Shot<br>Max Rounds: 1 | 0.714<br>(0.642, 0.771) | 0 (-1, 1) | < 0.001 | 0.543<br>(0.451, 0.624) | 0.739<br>(0.696, 0.783) |
| GPT-o3-mini Orchestrator<br>GPT-o3-mini as High-scoring Agent<br>GPT-o3-mini as Low-scoring Agent | Zero-Shot<br>Max Rounds: 1 | 0.768<br>(0.710, 0.814) | 0 (-1, 1) | 0.353 | 0.623<br>(0.542, 0.693) | 0.785<br>(0.749, 0.821) |

